# Does COVID-19 Testing Create More Cases? An Empirical Evidence on the Importance of Mass Testing During a Pandemic

**DOI:** 10.1101/2020.12.23.20248740

**Authors:** Ali Ahmed

## Abstract

The importance of testing and surveillance of an infectious disease cannot be underestimated. The testing is the first step to detect an infectious disease, and mass testing can slow or mitigate the spread of an infectious disease. Despite overwhelming evidence and the importance of testing discussed in the literature, there have been claims that “more COVID-19 testing creates more cases”. Therefore, there is a need to study whether massive testing is the reason for detecting more positive COVID-19 cases. In this research, we used a dataset from the U.S. Department of Health & Human Services and empirically showed that by increasing the COVID-19 testing in the U.S., the spread of the COVID-19 decreased significantly. Our results indicate a negative relationship between the number of positive cases and the number of tests performed in the past months. The large-scale testing may have helped identify positive and asymptomatic cases early in the course of illness, which enabled individuals to isolate themselves, thus reducing the chances of spreading the diseases and slowing the spread of the pandemic.

## Introduction

Detection and surveillance of an infectious disease is the first step in controlling a pandemic^1^. However, not much attention has been given to understand the role of testing in mitigating an infectious disease’s spread^2^. In this research, we present empirical evidence of how COVID-19 testing effects the spread of the disease and the number of positive cases in the future. We propose increasing the COVID-19 testing capacity in a given population may help identify asymptomatic and early cases, resulting in deterrence and decreased positive cases in the future. Specifically, we have analyzed a relationship between the number of positive cases reported in a month and the number of tests performed in the past months.

## Methods

We used a publicly available dataset released by the U.S. Department of Health & Human Services^3^. The dataset includes the COVID-19 laboratory test [Polymerase chain reaction (PCR)] results from over 1,000 U.S. laboratories and testing locations, including commercial and reference laboratories, public health laboratories, hospital laboratories, and other testing locations. The dataset contains the daily number of COVID-19 positive, negative, and inconclusive test results reported by each U.S. state’s health department. Using this dataset, we created a state-month panel data that includes the number of cumulative positive, negative, and inconclusive test results in each U.S. state for each month, ranging from March to November 2020.

To test the effect of COVID-19 testing on the number of positive cases in the coming months, we estimated two fixed-effects multiple linear regression models. Equation 1 analyzes the relationship between the number of tests conducted in the last month and the number of positive cases in the focal month. The main independent variable is the *LogTesting*_*i,t*−1_. The Equation 2 analyzes the relationship between the cumulative number of tests performed (*CumulativeTest*_*i,k*_) in all previous months and the number of positive cases reported in the focal month. The dependent variable for both equations is the log-transformed number of positive cases reported in the focal month *t* by the U.S. State *i*.

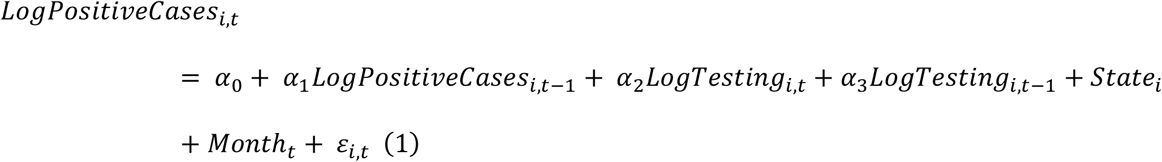

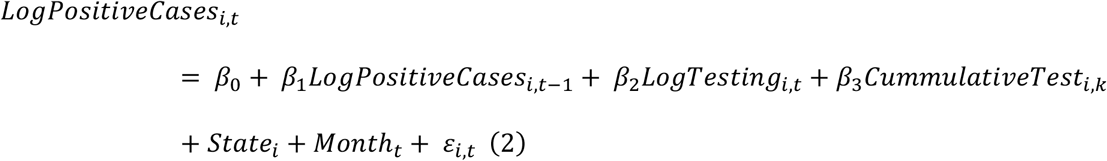

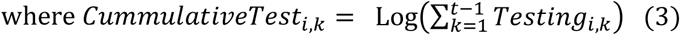

In Equation 1 – 3, *i* represents the 50 U.S. States, *t* indicates the month dummies on which the test results were conducted, ranging from April – November. The subscript *k* represents the lagged months, ranging from 1 – 10. The variable *State*_*i*_ is the fixed-effects controlling for the unobserved time-invariant characteristics of a U.S. state *i*. The variable *Month*_*t*_ is the time-dummy variable controlling for variations in disease spread from month to month. Finally, *ε*_*i,t*−*k*_ is the idiosyncratic error of the models.

## Results

Our results suggest a negative relationship between the number of positive cases and the number of tests performed in the past months. In Table 1, Column 1 and 2 shows the estimated models for Equation 1 and 2, respectively. Column 1 shows the negative effect of increasing the number of tests performed in the last month (*LogTesting*_*it*−1_) on the number of cases on the focal month within a State. Column 2 shows a similar pattern of decrease in positive cases by an increase in the number of tests performed in all the previous months (i.e. *CumulativeTest*_*i,k*_). The overall relationship suggests that having more tests in previous months reduces the number of positive cases in the coming months. Figure 1 shows the predicted relationship for both the models.

**Table 1:** Effect of Testing on Positive Cases in Future Months

|  | (1) | (2) |
| --- | --- | --- |
| | $LogPositiveCases_{i,t}$ | $LogPositiveCases_{i,t}$ |
| $LogPositive_{i,t-1}$ | 0.6337***<br>(0.04875) | 0.5962***<br>(0.04642) |
| $LogTesting_{i,t}$ | 1.1727***<br>(0.1146) | 1.0546***<br>(0.1052) |
| $LogTesting_{i,t-1}$ | **-0.6064\*\*\***<br>**(0.1096)** | -<br>- |
| $CummulativeTest_{i,t-1}$ | -<br>- | **-0.5595\*\*\***<br>**(0.09651)** |
| $Month_t = April$ | 0.4723**<br>(0.1897) | -0.8573**<br>(0.3506) |
| $Month_t = May$ | -0.4543***<br>(0.1353) | -1.5794***<br>(0.2678) |
| $Month_t = June$ | -0.2888***<br>(0.1059) | -1.2548***<br>(0.1901) |
| $Month_t = July$ | 0.03250<br>(0.07688) | -0.7411***<br>(0.1551) |
| $Month_t = August$ | -0.2606***<br>(0.07552) | -0.9515***<br>(0.1123) |
| $Month_t = September$ | -0.3222***<br>(0.06791) | -0.8028***<br>(0.09657) |
| $Month_t = October$ | -<br>- | -0.3574***<br>(0.1012) |
| $Month_t = November$ | 0.4031***<br>(0.1026) | -<br>- |
| $Constant$ | -2.9847***<br>(0.7316) | -0.9512<br>(0.7717) |
| State Fixed Effects | Yes | Yes |
| Observations | 430 | 430 |
| R-squared | 0.939 | 0.937 |

\*\* p<0.05, \*\*\* p<0.01

**Figure 1:**
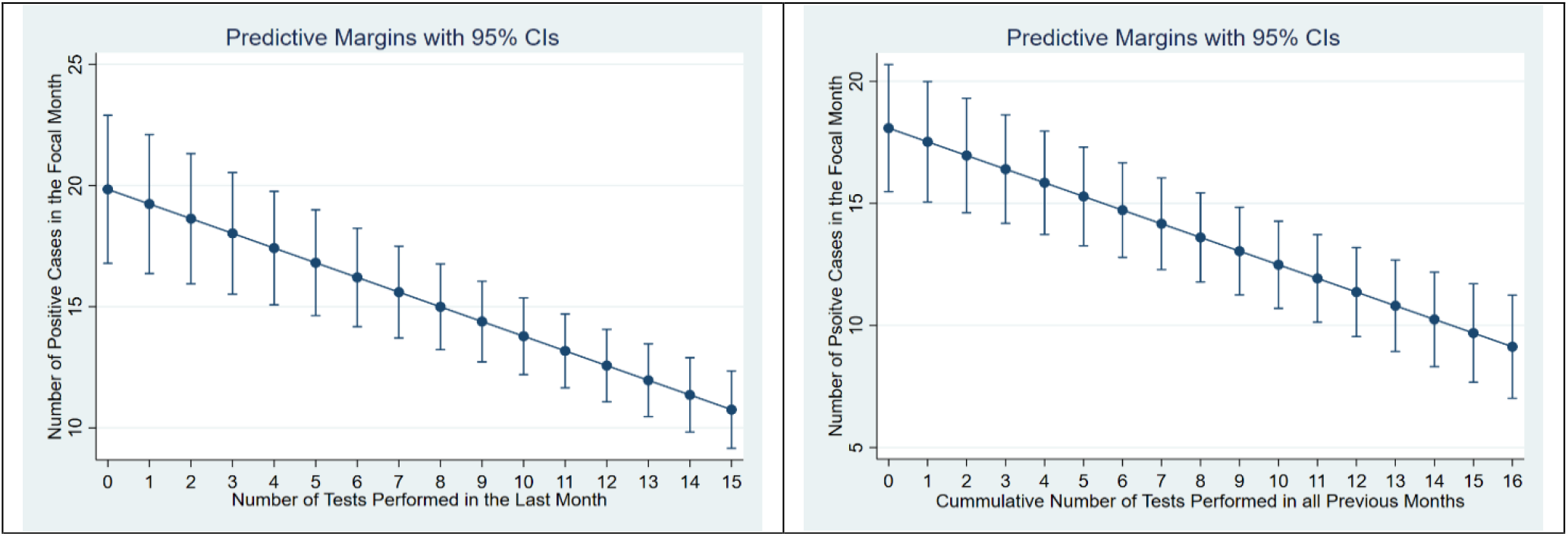
Relationship between the Number of Positive Cases and Tests in Previous Months

## Discussion

Our research explains the relationship between COVID-19 testing and positive cases. We provide empirical evidence on the importance of mass testing and its effects on controlling the pandemic. These analyses also refute the misperception that more tests yield more positive cases. We have shown that increasing the number of the tests yield fewer cases in the future—testing and surveillance help identify asymptomatic cases, which create deterrence and decrease the disease’s spread^2^. In short, mass testing mitigates the spread and yields less positive cases in the coming months.

## Data Availability

Department of Health & Human Services. COVID-19 Diagnostic Laboratory Testing (PCR Testing) Time Series. https://healthdata.gov/dataset/covid-19-diagnostic-laboratory-testing-pcr-testing-time-series. Published 2020.

## References

1. Manabe YC, Sharfstein JS, Armstrong K. The Need for More and Better Testing for COVID-19. JAMA. November 2020. doi:10.1001/jama.2020.21694

2. Augustine R, Das S, Hasan A, et al. Rapid Antibody-Based COVID-19 Mass Surveillance: Relevance, Challenges, and Prospects in a Pandemic and Post-Pandemic World. J Clin Med. 2020;9(10):3372.

3. Department of Health & Human Services. COVID-19 Diagnostic Laboratory Testing (PCR Testing) Time Series. https://healthdata.gov/dataset/covid-19-diagnostic-laboratory-testing-pcr-testing-time-series. Published 2020.

